# Altered smell and taste: anosmia, parosmia and the impact of long Covid-19

**DOI:** 10.1101/2020.11.26.20239152

**Authors:** Duika L Burges Watson, Miglena Campbell, Claire Hopkins, Barry Smith, Chris Kelly, Vincent Deary

## Abstract

**Background:** Qualitative olfactory (smell) dysfunctions are a common side effect of post-viral illness and known to impact quality of life and health status. Evidence is emerging that taste and smell loss are common symptoms of Covid-19 that may emerge and persist long after initial infection. The aim of the present study was to document the impact of post Covid-19 alterations to taste and smell.

**Methods:** We conducted exploratory thematic analysis of user-generated text from 9000 users of the AbScent Covid-19 Smell and Taste Loss moderated Facebook support group from March 24 to 30th September 2020.

**Results:** Participants reported difficulty explaining and managing an altered sense of taste and smell; a lack of interpersonal and professional explanation or support; altered eating; appetite loss, weight change; loss of pleasure in food, eating and social engagement; altered intimacy and an altered relationship to self and others.

**Conclusions:** Our findings suggest altered taste and smell with Covid-19 may lead to severe disruption to daily living that impacts on psychological well-being, physical health, relationships and sense of self. More specifically, participants reported impacts that related to reduced desire and ability to eat and prepare food; weight gain, weight loss and nutritional insufficiency; emotional wellbeing; professional practice; intimacy and social bonding; and the disruption of people’s sense of reality and themselves. Our findings should inform further research and suggest areas for the training, assessment and treatment practices of health care professionals working with long Covid.

## Introduction

Qualitative olfactory dysfunctions after viral infections have been recognised as affecting mood, food enjoyment, reducing ability to detect dangers, influencing health status and impacting on social life(1, 2). Moreover, in reports of post-viral smell alteration, studies have found that as many as 56% of patients experience parosmia (distorted smell in the presence of a familiar odour source) and phantosmia (experience of smell in the absence of an odour source)(3). These are recognised as having a particularly pronounced impact on quality of life as most experiences involve unpleasant smells (malodours)(4). These distortions of smell have a profound impact on our experience of eating and enjoyment of food since what we typically regard as the taste of what we are eating is actually due to the combination of smell and taste: odours released when we chew foods or sip drinks combine with the basic tastes from the tongue (salt, sweet, sour, bitter, umami) to create the unified experience of flavour. Therefore, loss or distortion of the sense of smell leads to loss of or distortion to our perceptions of flavour, commonly described as tastes. Although evidence is still emerging about the nature and impact of smell and taste loss following COVID-19 early self-reports on social and mainstream media indicate that it is having a considerable impact.

It has become apparent that alteration to smell and taste is one of the most prevalent symptoms of COVID-19 (5). However, while the governing body for Ear Nose and Throat specialists in the UK had recognised that loss of smell and taste could be used to predict COVID-19 status as early as 24 March(6), leading health institutions and public bodies were slow to acknowledge it or to provide this information to the public (7). It took until 18 May until the U.K. added loss of smell and taste to the list of official recognised symptoms. Moreover, the focus on predicting COVID-19 status due to concerns about transmission eclipsed attention to the *impact* of smell and taste loss. Although a large proportion of people recovered smell and taste within weeks, around 10% reported persistent problems including anosmia (loss of smell), hyposmia (reduced smell), parosmia and phantasomia as well as dysgeusia (distortion of basic tastes such as salt, sweet, sour, bitter) and reduced chemesthesis (chemical sensitivity experienced as sensations such as the burn of chilli, cooling of mint or warmth of ginger) (8)(Parma et al, 2020). Given the steady rise in numbers of COVID-19 infections globally, there are potentially millions living with altered sensory experience. However, little is known about its impact, and despite increased recognition of long covid and long-haul impacts, alterations in taste, smell and chemesthesis are still being overlooked by even the most well-informed health practitioners (9).

The neglect of our chemical senses is not new; their alteration in long term health complaints such as Parkinson’s disease, acquired brain injury and cancer has often been overlooked. There are few helpful interventions and a relative lack of interest in food hedonics or food related quality of life issues (10, 11). As a result of the relative neglect of olfactory and gustatory dysfunction, and lack of concern in the medical world for the emotional impact of altered eating and smell distortions, people are seeking help and support outside of medical care settings. The UK charity AbScent, ENT UK and the British Rhinological Society (BRS) collaborated in developing a range of resources to meet this need. AbScent established the COVID-19 Smell and Taste Loss Facebook group to meet the growing demand for information and support. This website has gathered international media attention as a space where people could

> “read about the experience of others and connect with olfaction experts via online seminars and presentations.” (12).

With the collaboration of the founder of this online community and the active participation of its users, we have set out to address the question: *what is the impact of post COVID-19 alterations to our senses of smell and taste?* To date little is known about the psychosocial impact of olfactory and gustatory changes related to Covid-19 (see Leopold 2002). As such this constitutes exploratory research which we hope will provide a foundation for future exploration of these important issues.

## Methods

### Ethical Considerations

The ethics of digital social research are an evolving issue, with new guidelines continuing to emerge from different statutory and regulative bodies. More recent guidelines such as those of the British Psychological Society (BPS; 2017) offer principles for consideration focussing on these key domains: the public-private domain distinction online; confidentiality and security of online data; procedures for obtaining valid consent; and implications for scientific value and potential harm. Similarly, the British Sociological Association’s Ethics Guidelines and Resources for Digital Research suggest that the ethics of any given project will be in part specific to the context in which they are conducted.:

> Each research situation is unique and it will not be possible simply to apply a standard template in order to guarantee ethical practice. Rather, we should consider the situational ethics of digital research, taking very carefully into account the context and the implications of conducting this research
>
> BSA (2017)

Following consideration of these guidelines the following ethical issues were identified in this research, and the following ethical principles were adopted.

With regards to consent and the private/public distinction, the AbScent group is a closed group in which participants once admitted can freely exchange information. Before admittance they are presented with information about the site that makes it clear that there are researchers in the group who monitor the exchanges for potential research themes. However, none of the posts that were part of the spontaneous and unprompted activity of the group were used in the thematic analysis of this paper. Once we deployed clearly research-driven question prompts to the group (see methods below) it was made clear that any answers to these research prompts were for research purposes and might be anonymised and used in a future research output. In addition, once we had identified representative quotes for inclusion in the research paper, we contacted each person individually to ensure their consent for their anonymised quotes to be included. All provided consent. Further, once an initial draft of the paper was complete, it was shared on the closed group for comment and emendation. This generated hundreds of entirely positive comments, including several participants expressing “tears of joy and relief” to see the issues they were dealing with getting wider attention. As such we believe that with regard to scientific value and potential harm, the latter is minimal, the former considerable. We also, in line with the guidance of the BSA (2017) invited those who had substantially quoted or commented to be co-authors. All declined, but were happy for the work to be published with their quotes included. To protect participant anonymity and confidentiality of the remaining data set (of unused quotes) we have withheld this data from wider availability. This is because of the potential for unquoted items within the data set to be back traced via internet search engines. This research was granted ethical approval by the Newcastle University Ethics Committee (Ref: 3058/2020).

#### Research Team

CK is the founder member off AbScent, and is a former anosmic and parosmic. DBW, a health geographer, also acted as a moderator of the group. The other authors consist of health psychologists (VD and MC); an ENT specialist (CH); a philosopher and sensory expert (BCS).

#### Study Design-setting

The *AbScent Covid-19 Smell and Taste Loss* Facebook group was formed in March 2020 and by September 2020 had over 9000 active members. Membership requests were screened by a series of entry questions that ensure the focus is on smell and taste issues and not broader COVID-19 complications. Posts to the group are monitored for topicality. Every new member was welcomed in a group post with detail of the information resources available, group rules and stories in the recent threads. The majority of members were from the UK (43%) and USA (26%) with smaller contributions (3% or less) from South Africa, Philippines, Sweden, Pakistan, India, France, Nigeria, the Netherlands and elsewhere. Individuals’ use of the site varied with some continuing regularly to post updates of their experience from March – September, whilst others posted only during the initial period of their infection, or when there was a new symptom or change in symptoms. To our knowledge, all members were 18 or over. As this was exploratory and novel research, we did not screen or choose comments on the basis of gender or age range. Moderators also posted links to relevant news items, research articles, information sources, events and survey/research questions and links on a regular basis. New membership and daily comments continue at time of writing.

#### Study Design-data collection and analysis

There were two distinct phases to this project; a pre-research phase and the research phase. In the initial phase, the founder and moderator of the group (CK and DBW) managed and administered the group, and also provided access to resources for participants. During the conduct of this work, they noticed recurring issues emerging amongst the rapidly growing membership. This led to the research phase of the project where the research team were assembled by CK and DBW and together they agreed upon a prompt question which was put to the online group for the explicit purpose of generating research data on the impact of smell and taste alteration in long covid. This was the prompt question:

- Detail your personal journey with smell loss and its impacts

It generated over 100 individual posts (from a paragraph to multiple paragraphs, with multiple chains of communication for each post). These were thematically analysed by DBW, CK and MC. All co-authors then discussed and agreed on these themes. Next these themes were used to generate 3 follow up questions:

- Describe changes in experience of body odour: (595 individual posts; from a sentence to several paragraphs with multiple chains of communication for each post)
- How has changed smell affected your intimate relationships? (91 individual posts from a sentence to several paragraphs with multiple chains of communication for each post)
- Can you tell us a bit about how your relationship to food has changed? Has this affected what you eat and how much you eat? Has this had an impact on your body and/or body image? (213 individual posts; from a paragraph to multiple paragraphs with multiple chains of communication for each post)

These were posted to the AbScent Covid-19 Smell and Taste Loss Facebook site from September as moderator announcements. Again CK, DBW and MC thematically analysed the themes which were then refined with the other authors. Participants were also invited to comment on themes and to contribute any further themes or ideas they felt important to living with smell and taste disturbance and Covid-19. All efforts were made co-produce this research (10, 15). On-line discussions between the CK, DBW and contributors to the group were used to check each of the themes with members, and in each instance to offer them the opportunity to discuss, refine or correct any misunderstandings. For each announcement we stressed that anonymous quotes would be used in the research. For each quote used in the paper we also re-checked individually that participants consented to their words being used in this research paper. Finally, the pre-print of this paper was posted for comment and emendation, as outlined in the ethics section above.

## Results

In the following sections, we begin by discussing how participants first presented their altered experience in the FB group, how they engaged with one another and began to make sense of their experience. We go on to describe the consequences for participants of altered eating before going on to describe patients’ reports of altered experience of the world, of their relations to one another and to themselves, before moving to a discussion of these findings.

Three broad themes were identified

### 1. Making sense of altered experience: “we are the research”

New members of the FB group experiencing sudden onset anosmia posted throughout April to September and repeatedly stressed a difficulty in understanding and managing smell and taste loss, particularly in the absence of support from health professionals and t[he people closest to them] significant others. Making sense of their altered experience was an emotional and cognitive burden for participants; a task that comprised three main components.

#### 1.1 A long, unique and unpredictable journey

The only constant in the course of smell and taste alteration was its sudden onset:

> “It was like a light switch: from 100% to 0% in a couple of hours…No distorted smells, no whiffs, nothing. It’s like my nose is switched off
>
> My taste and smell left on the Sunday, very suddenly. One minute I was eating, the next it had gone.”

From there on, comments reflected the ongoing challenge of the condition that for most kept shifting unpredictably. Common expressions included “what the hell is going on?”, “can anyone explain?”, “why could you eat eggs one day and not the next?”. This applied not only to shifts in taste and smell but to the overall experience of the post COVID-19 self:

> “It’s scary to just not be sure of what is going to happen next with my post COVID body… it’s a new symptom/feeling every week it seems.”

Posts from group members provided detailed narratives of the changes in their sensory experience of smelling and tasting. These typically highlighted shifts in the experience through different phases that were likely to include some or all of the following: anosmia and taste changes; fluctuating/minimal returns of taste and smell; hyposmia; parosmia; fluctuating parosmia; “new normals” of stable but ongoing distortions, and/or recoveries. Group members’ accounts indicated ongoing confusion and frustration, particularly given other participants experiencing quite different trajectories. As many posts reveal, not everyone was ‘going through the same thing’ and not always at the same time. Even narratives that charted recovery were frequently not smooth and straightforward. For instance, one person posted in April after 11 weeks of no smell and taste. In early May they reported some return of taste and flavour which was short lived; later that month posting that there is now a ‘burning sort of taste and smell’ then, later that month, an ‘awful metallic taste’. By June they posted a continued but less intense metallic taste mixed with ‘some parosmia’, then in July with an easing of metallic taste and parosmia, some improvement associated with smell training but still fluctuating and distorted. In early August feeling they had reached a ‘plateau’, still with distortions in smell and taste but less intensity.

For others there was much less change, and the uncertainty of ‘recovery’ was more evident:

> “I’m mostly worried about the complete lack of any progress. Even distorted smells and tastes would be welcome”

Day to day variation in symptoms was also commonly reported:

> “There are good days and bad days now… Some days something won’t smell bad, but the next day it does.”

This lack of both short- and long-term predictability, and the singularity of the reported experience, was captured by one participant thus:

> “I think it’s really important for us to partake in these research projects because we are the research. There is no one before us. For those of us who have been dealing with this for over 5 months, we are the longest known cases of this craziness that no one really has answers for.”

This sense of being pioneers of a new phenomenon which had little existent help or understanding was the next theme that emerged from the analysis.

#### 1.2 No help or understanding

> “It’s so difficult to describe it to people that haven’t experienced it!
>
> It’s been hard for people even close to me to understand the severity of the loss and how it’s affected my life.”

Part of the cognitive and emotional labour of coming to terms with sensory change, was how difficult it was for others to understand it. Explaining the impact of smell and taste loss to significant others was noted by many as a challenging issue. This was in part because smell and taste loss are an invisible condition, i.e. unless there is dramatic weight change, there may be no visible signs of its existence. Many people described their attempts to explain their lived experience as being met by claims of exaggeration, “being dramatic”, or other forms of minimisation. This included jokes, suggestions that the inability to register bad smells was an advantage; “you might lose weight”, “you were lucky to be alive”. Leading a thread of communication on this topic in September one person posts:

> “When I tell someone I have lost both taste and smell, they react as though it were something minor. It’s 2 of the 5 senses!! People seem to show very little interest or sympathy. I realize it’s not as horrible as going deaf or blind. I would just expect more than a blasé response. Is it just me?”

Hundreds of comments follow that confirmed that lack of empathy, understanding and support from others was a common experience. Those commenting reflected on the downward comparison with other sensory losses such as hearing or sight, or on their own lack of attention to smell and taste “until it was gone”. This was often used as an attempt to ameliorate and explain the dismissive attitude of loved ones: “they can’t understand unless they experience it themselves”. Others explained they no longer mention the sensory impacts, because it was too difficult to explain, they felt guilty doing so, they found it too upsetting to even talk about, or perceived no one cared:

> “I’m sick of myself complaining.”
>
> “Sometimes this thing occupies my mind so much I just want to tell people about it, to try to compare what I smell/taste with what they do, and just to express how it feels to hate things that I used to love, but they don’t want to hear it.”

This lack of help and understanding was also an issue when it came to health care professionals. A moderator-added research question asked participants to comment on what they would like from the healthcare system. The most common answer was more understanding and empathy, particularly in recognising such symptoms are “severe” and not to be dismissed. People reported feeling “abandoned” and having a range of unhelpful responses from clinicians including a GP who had told them to “come back in six months”, another GP being “baffled”, yet others who were unable to offer any explanation or empathy; ENTs that offered “no help”; a dentist suspecting it was an allergy. Participants perceived their problems as being out of reach of current health expertise:

> “It feels like it will be years before medical knowledge catches up to what we are experiencing. And that is a strange place to be.”

However, what was also clear over the course of time is that people did begin to reach an accommodation with their symptoms, and formed a language for understanding and describing them to self and others. This was often done through peer support and through dint of prolonged experience. This hard-won shift from lack of understanding to being able to comprehend and manage their condition, and share expertise with others, was the last component of their making sense of the conditions they found themselves in.

#### 1.3 From chaos to quest

The sociologist of health and illness Arthur Frank describes in his book “The Wounded Storyteller” (16) the narrative work of having an altered state of health. He distinguishes between “quest narratives” and “chaos narratives”. The latter are typified by the kind of processes described above where neither the individual, nor their significant others or health professionals, can make sense of their condition. This state is marked by the kind of isolation and distress evidenced above. Quest narratives, by contrast, enable the person to make sense of, and derive meaning from, their altered state. This is enabled, Frank argues, by hearing other stories that echo theirs, this in turn enables them to tell their story for themselves and others. This work of mutual making sense was very much in evidence in the Facebook community. This was often evident from first posts, where participants frequently expressed a sense of relief at finding the site; that they were “not alone”; that they “had not realised till now that others were going through the same thing”.

Part of what enabled them to make sense of their experience was finding a shared language. Our data suggested that finding the right medical and scientific labels and explanations validated, legitimised and normalised people’s experiences and allowed them to narrate and manage their experience better.

> “A knowledge of the terms has been immensely valuable for me. Parosmia in particular is very hard to describe, and other people find it hard to believe and easy to dismiss, so referring to a recognised condition helps me to tell other people about it. Also, for my own piece of mind, before I understood what was wrong I was scared and confused. Although I still don’t really understand the conditions now, I now feel I have enough knowledge to keep things in perspective. The more I learn, the more I feel in control.”

By September, many people posting to the site had experienced smell and taste changes lasting more than half a year. More chaotic narratives appeared now somewhat easier to narrate, with trajectories that included more recognisable patterns over the long term. A common thread was failure to find help in conventional health settings and only feeling understood within the supportive environment of the (moderated) Facebook group.

There was also some evidence of new terms being coined. People struggled with overwhelming and often pervasively unpleasant parosmic smells. They were described with words that included: sewer, cat food, spicy, pungent, strong herbs, sickly metallic, dirty fish tank, off milk or yoghurt, sweet and grassy, dog food, curry, garlic, sickly sweet metallic, kippers, chemicals and fruity sewage. Over time, attempts to describe these “indescribable” smells were replaced with a shared shorthand of “the Covid smell”.

There was also evidence of people adjusting to their condition and finding ways of managing it. Again, this was often perceived as being mediated by the group:

> “This group has helped me immensely to break out of that mindset of ‘I’m not going to eat because everything tastes terrible’ by seeing what others are finding tolerable, giving me new things that seem safe to try”

This determination to continue to experiment often resulted in new knowledge about how to “work around” smell and taste deficits:

> “It was bad at the beginning, but I’ve adjusted to foods I can eat and can’t eat now. Some things I just ploughed through and tried to get used to the adjusted taste. Some things still make me feel sick, like washing up liquid and perfume, but most things I can cope with.”
>
> “I can still cook some things which give me joy. And I’m trying to focus on some other aspects of the ingesting experience like temperature and texture. I am relying a lot on the comforting aspect of soup, the surprising aspects of salads and crunchy snacks”

This ongoing accumulation of terms, encouragement, hints and tips for coping in turn seemed to help others make sense of their condition:

> “Thank goodness for this group!”

### 2. Altered eating and its consequences

Besides the work of understanding, explaining and managing their condition, the most direct impact reported was on the ingestion and enjoyment of food. This was mediated by a profound alteration in the smell and flavour of food, mainly due to the role smell plays in the perception of flavour, which most people think of as *taste*. Again, there was not one pattern here, but there were some repeated themes around altered flavour perception and how it linked to weight loss, weight gain, pleasure and socialising. The impact on health, weight and body image was most commonly described as negative, but not exclusively. Further, many people recounted different sensory changes at different stages of the Covid-19 journey, with concomitantly variable effects on weight loss, gain and nutritional compromise.

#### 2.1 Appetite and weight changes

Most participants described anosmia and the concomitant flavour changes as having major impacts on appetite, enjoyment, fullness and satiety. Food became bland and unappetising resulting in a reduced desire to eat, cook or participate in food related activities. Parosmia and phantosmia had even more harrowing effects on food and eating. People reported pervasive “off” smells, or a metallic taste:

> “All food tastes/smells just too disgusting to eat. Only yoghurt is ok. So I lose weight instead of gaining it.”
>
> “Everything tastes awful for me as well, so you are not alone. Makes me not want to eat.”

The effect of these flavour changes on diet and diet quality and content varied greatly from person to person. Anosmia could both result in a diet of more sugary, fatty, highly processed foods because “processed food tastes the least offensive” (-since basic tastes like salt and sugar, detected by the tongue, were still perceptible) or for some (at least initially) an improved diet away from unhealthy foods that had previously been consumed for their flavour. Both with anosmia and parosmia people could be fearful about eating unsafe foods. Universally, parosmia resulted in a much-reduced selection of foods that almost always raised concerns about health. Overall this experience of altered flavour was reported as pushing appetite and intake in one of two ways.

For some participants flavour loss shifted preferences towards increasing food intake as it “takes more to hit the spot”. Eating now involved chasing high impact taste and trigeminal sensations such as sugar, salt and piquancy. The most commonly described foods included crisps, chocolate, chilli crisps, and other items that provided unusual textural experiences. For some this increase in consumption of snacks resulted in a reduction of intake at mealtimes. But others reported becoming “insatiable”, “always hungry” and even losing control around food:

> “food satisfaction is lacking and I see myself eating more to try to get that satisfied feeling. My weight has gone up. Just one more depressing reminder of this illness. I am gaining weight due to a constant urge to satisfy what can never be satisfied”

For others flavours had become so unpleasant that food was avoided leading to weight loss as well as other cognitive and emotional consequences:

> “Four months into recovery the rancid/metallic taste and smell hit me. Since then I cannot eat much… I simply can’t eat enough to workout AND my shortness of breath didn’t help. Since then, I’ve been rapidly losing weight and I honestly do not recognize my body. This affects me mentally because I don’t like the way I’m starting to look. This has been the hardest thing I’ve had to go through. I dread eating and even going to restaurants or being around food is hard for me. It’s disgusting. I have to force feed myself to get nutrients and honestly I’ve lost my appetite because of how nasty food tastes and smells. I have absolutely no energy and severe fatigue. My eyes are sunken in from malnutrition.”

Weight loss was commonly reported, but this was not always considered to be a problem:

> “I’ve lost weight since I don’t want to eat, I don’t have a desire to eat, very few cravings. I eat mostly because my body tells me I need to. I don’t mind that I’ve lost weight. I’ve been trying to eat healthier. More vegetables. I figure, What’s the point of eating much else since it all tastes the same.”

#### 2.2 Pleasure and commensality

Food and eating it with other people is a major source of daily pleasure and social bonding that is often not appreciated until it is no longer possible (17, 18). Research into altered eating in head and neck cancer survivors has shown that commensality - the social sharing of food - was one of the most grieved aspects of an altered relationship to food (10)(Burges Watson et al, 2018). This was a prominent theme for the post COVID group too:

> “I am grieving for my lost senses. No more wine and cheese tasting nights or gin cocktails with my ‘girls’ - the only alcohol I can manage is vodka and orange!”

The wider impacts of altered eating were visible in many aspects of people’s quality of life. Posts revealed loss of food related joy and pleasure and the associated joy in anticipation of food.

> “Eating used to be a pleasure, something to look forward to. Now food is just sustenance…As there is no pleasure in eating, I stop as soon as I am full. And with everything tasting bland, I find that I feel full very quickly… I find it extremely depressing.”
>
> “The joy I had had disappeared. All I can comment on in the texture of the food I am eating. I can tell if something is sweet or savoury and my tongue tingles if it’s spicy but I have zero definition of specific flavour. I have almost no appetite and eat because I know I have to in order to stay healthy. I get no enjoyment from my food anymore and the only thing I seem to enjoy eating are cold, peeled apples because I like the refreshing sensation in my mouth.”

Participants’ family and social interaction was diminished too, with participants reporting that they no longer could cook for their family, that they had to leave the house if cooking was taking place, and a host of other social consequences: “I feel very not fun to share meals with”, “repetitive diets because only some things edible”, “can no longer go out to eat because of the smell of the venue”, “can no longer go out for a coffee with friends”, “loss of pride in being a ‘non-picky’ eater”. Several commented on the perceived effect on family life:

> “My relations with my family are strained, or that’s how I perceive them anyways… My husband doesn’t seem to understand how devastating it is to lose these senses, although he tries his best, I do believe that.”

### 3. Altered relationship to the world

Much more than the relationship to food was altered by COVID-19 associated sensory changes. Smell serves to orientate us to our environment, to other people and places, and to ourselves, in terms of comfort/familiarity and as a signal of novelty/threat. The disruption of these cues was reported as fundamentally changing the relationship to the world, self, and others in a way that was often described in terms of a fundamental existential upheaval or assault:

> “The world is very blank. Or if not blank, shades of decay. I feel alien from myself. It’s also kind of a loneliness in the world. Like a part of me is missing as I can no longer smell and experience the emotions of everyday basic living. Detached from normality. Lonely in my body. It’s so hard to explain.”

#### 3.1 The world of things

This profound disorientation can be traced in part to the uncoupling of objects, their associated smells and the emotions to which these normally give rise. Things not smelling at all was reported as inducing feelings of detachment, dissociation and unreality:

> “You feel so detached from reality when you can’t smell your surroundings.”
>
> “I feel discombobulated - like I don’t exist. I can’t smell my house and feel at home. I can’t smell fresh air or grass when I go out. I can’t smell the rain. I would say I am mildly depressed about it and cry sometimes”

At the other end of this spectrum, a “covid smell” or smells, i.e. assorted unpleasant parosmias and phanstosmias, could eclipse other sensory experiences. The intensity of parosmic smells and/or persistent unpleasant tastes could elicit a “gag” response, cause people to vomit and they were frequently persistent and inescapable:

> “Worried if you get a passing smell as you know it’s going to sit in your nose all day. The smell in the car was so horrible! Before I could hardly wait to be home to eat a slice of new made pizza. Now I had to drive with full aircon and open windows. I was on the edge of what I can take.”

This disorientation was also attributable to odours being “misclassified”. Things no longer smelled like they should, and food became confused with non-food in unpredictable and upsetting ways:

> “Wine smells like sewage. Prosecco is even worse.”
>
> “I’m getting horrible smells for most things that don’t fit with what they are”

These sensory confusions were reported as “unbearable” for some, with everything smelling burned, of ash, or food smelling “chemically”, or “like the fridge”, or of “single use plastic”, or “cleaning products”. There was a long thread on “poo smells” which evinced some of these issues around sensory mis-categorisation. Poo was described as smelling of “cat food”, a “sweet fruity smell”, of “rancid wet hay” or a “sickly smell like eggs”. Troublingly for some, this was now a similar smell to food and drinks like “onions” and “coffee”. Indeed, whilst food had become more subjectively disgusting, poo was frequently described as becoming a less repulsive smell or as having a more “faint Covid smell” than some food stuffs. As one person summarised it:

> “Poo now smelled better than coffee.”

A discussion thread between the researchers and two participants provided a particularly noteworthy example of how faecal smells troubled the boundaries with food, making it difficult to distinguish the edible from the toxic; as faeces now smelt “so close to the new parosmic distortion of food”:

> “then, when I got the same [poo] smell from my coffee, hot toast etc it was vile. I’m not sure if it’s because I now associate it with poo or because your brain tells you that you shouldn’t be eating things that don’t smell right.”

The effect of this was also for some felt in their professional life. For those who partly relied on their nose to do their job (e.g. nurse, food writer) an essential tool had gone missing rendering them professionally less effective:

> “I can’t detect faeces, urine, blood, infected wounds, flatus or any bodily functions. As a nurse, I’ve lost one of my most important tools - my sense of smell often tells me more than I can see. Professionally, anosmia makes things so much more difficult.”

#### 3.2 Altered intimacy: the world of self and others

As well as the things in the world, people also reported their social world was impacted by taste and smell alteration:

> “I miss smelling things like fresh cut grass, clean laundry, and the scent of my significant other.”

There was a long discussion of the relationship between smell and intimate and sexual relationships, with a frank acknowledgment of the importance of the smell of the romantic/sexual partner in close relationships and physical intimacy. Body odour is known to be significant to sexual relationships in facilitating the detection of key factors that signal compatibility, maintaining familiarity and security within a relationship, and moderating sexual desire(19). Not being able to smell your partner was a recurring and dominant theme throughout this discussion, with many people missing the smell or “unique scent” of their partner. This could also switch to active aversion and disgust with parosmia:

> “I can’t smell my boyfriend’s natural scent, which makes me feel more distant from him. Like he is a stranger. I used to feel comforted being able to smell him while cuddling. Worse is that his kisses taste really bad to me now, so I avoid that, but haven’t told him because I don’t want to hurt his feelings. Also I am constantly worried that I smell bad myself and it makes me very insecure.”
>
> “The worst bit is not knowing if I smell. It makes me really self-conscious. If we get intimate I can’t get lost in the moment anymore because I’m constantly thinking ‘what if I stink? Is that roadkill smell me or him?’”
>
> “His natural odour used to make me want him; now it makes me vom. I can’t tell him. Imagine your partner saying that to you?”

For those that did report talking about it, the result was not always a better understanding. The following long quote details the journey from a relatively unaffected sex life with anosmia, to much greater difficulties in intimacy and understanding brought on by parosmia:

> “The new problem was his rotten breath. It was unbearable, no matter how hard I tried to put it out of my mind and make do. The first few times I turned away, he accused me of exaggerating, and he insisted that there was no possible way his breath was a problem. It made me feel stupid. I wished so badly that I was making it up. I wished it were a figment of my imagination, a dramatic charade I could humbly admit to falsifying and then move on, put it behind us. But it was abhorrent. It still is. Between us, there is this wall that says ‘Stop! Too far. You cannot pass this line’. We have to play this odd game of fumbling around in bed to make sure the foul odor in question does not make its way to my defective nose and throw sex out the window entirely.”

However, there was also another side to this. One commentator reported better intimate relationships and a more adventurous sex life as the result of their anosmia:

> “I don’t miss human smells, good or bad, at all. It makes for better connections with people because I’m not offended by their breath or smell…Loss of smell means I am much less inhibited. I used to be so sensitive to smells and that would get in the way of how much or how little I participated. Now that I really can’t smell much I’m much more willing to do different things. It’s greatly improved my involvement in my intimate relationship participation.”

It was not only romantic and sexual relationships that were affected. A few comments also addressed the impact on maternal bonding with babies and children, and the impact on dating for those looking for a relationship, issues that had come up frequently in our passive thematic analysis.

> “a lot of my maternal bonding feelings for my children are tied up with smell I’m single but avoiding dating as I can’t judge my own body smell accurately, and can’t imagine what anyone else smells like! so I’m preferring to avoid knowing”

This last quote highlights the problems of an altered relationship to self, brought on by smell loss or distortion, as also evinced by some of the quotes above. Several commentators pointed out the effects of loss of self-smells, often called in the discussions “body odour” but not with the negative connotation this term often has. Body odours in the discussion could refer to unadulterated “natural” smells of the body and added smells from things such as soaps, deodorisers and perfumes. Body odours were of particular concern both in terms of relationship to self and others, and because many commented that the loss of ability to register natural body odour was the first thing they noticed, and the last thing to return. Participants mentioned armpits, menstrual blood, faeces, urine, farts and sweat as casualties in their altered relationship to their own body odour. With these smells either absent or distorted by parosmia, many participants lived constantly on in fear of being anti-socially “smelly”. There was frequent use of the term “paranoia”, and multiple reports of frequent showering, multiple changes of clothing throughout the day, deodorising and constant cleaning, almost to the point of obsession according to some.

> “Personally, I can’t tell if I have BO (bordering on paranoia)”
>
> “I’m worrying because a) I don’t know if I smell in some way b) I don’t know if my house or toilet smell c) had a couple of urine infections in the past and a strong smell was the first sign”
>
> “At first, I couldn’t smell any sort of bodily functions at all. Now that I’m in the parosmia stage, the best description I can give is it all smells like sweet wet hay. I smell it strongly regularly, and it has made me paranoid that I smell bad to others even when I logically know I don’t.”

Apart for the importance of self-smells to the perception of others, some also commented on the importance of it to the sense of self:

> “I’ve found that not smelling like ‘myself’ has had a great impact on me. I am so used to my own smell, that smelling something else is alien, I don’t feel like I am me anymore.”

## Discussion

This exploratory co-produced research has brought to light the impact of smell and taste loss and distortion on a large cohort of self-reported COVID-19 Facebook members. (We continue to refer to ‘smell and taste loss’ although we are mostly talking about smell loss and its impact on what people call taste, i.e. perception of flavours). The impact is extensive. At the broadest level, the impact shows up in the work of having to manage a poorly understood and fluctuating condition; in an altered relationship with food that includes loss of pleasure and changes in appetite and weight; and in an altered relationship to the self, the world and other people. Many of these impacts in turn negatively impacted upon mental health.

People with smell and taste loss or distortion may have the burden of living with an invisible illness, which nevertheless leads to severe disruption of daily life and routines. This combination of invisibility and disruption means that those with smell and taste loss can be held “accountable” in the sense meant by Garfinkel (20). They breach the “seen but unnoticed” rules of daily living, the “ceremonial order” of food, eating and intimacy, but do so for no obvious or visible reason. As such, part of the work of having smell and taste loss can entail having to constantly account for oneself and one’s experiences to an often uncomprehending audience of family, friends, colleagues and health professionals. This is in addition to the cognitive, behavioural and emotional labour occasioned by attempts to come to terms with and communicate what it’s like to live with smell and taste loss or distortion.

The Altered Eating Framework (10) identifies that in an altered relationship with food, multiple domains of life can be affected. This is evinced by this work where in addition to the interpersonal and cognitive labour of trying to understand and explain the condition to self and others, there were real, often worrying, physiological consequences in terms of weight loss, weight gain and malnutrition; there were profound disruptions to social, family and love lives; and an altered relationship to the world and the self. Despite some exceptions, most of these impacts were negative.

The altered eating framework suggests that the emotional domain of altered eating problems be seen, in part, as the cumulative tally of the impact of the issues with food on fundamental well-being. With COVID related smell and taste loss/distortion, people reported a range of emotional responses including worried, happy, disgusted, confused, frustrated, depressed, anxious, and hopeless. Some of these consequences were profound:

> “I’m losing hope and I’ve never been more depressed in my life. Will I ever get better? This has left me so low in mood. It’s really quite debilitating - physically, mentally and professionally. I’m 6 months in and losing hope.”
>
> “I have noticed a definite shift in my mood since all this started. When I look in the mirror there is definitely a slight sadness and emptiness in my eyes that was never there before. I think I am feeling a little disconnected from my normal place in the world…”

Of course, some of these comments may be attributed to the broader impact of COVID-19 and post viral sequalae, though the group did tend to stay focussed on smell and taste loss and its impact.

Previous medical and nutritional literature on smell and taste loss has not always brought to light this broad range of profound consequences, focussing more on how diet choices may change, but rarely is an association made with nutrient deficiency. The literature tends to suggests most people maintain “adequate dietary intake” (2). This was not the case for many of our participants, with widespread reports of weight gain, weight loss and malnutrition, and consequences that went far beyond the nutritional. In August the BRS issued consensus guidelines for treatment of smell and taste loss, noting the surge in cases that would need to be managed at the level of primary care (21). Our research would suggest that this guidance be extended to other professionals and that it contains an awareness of the extent and seriousness of the broader psychosocial impact of smell and taste loss. To date there is little input on this in the training curriculum of GPs, dieticians, nutritionists and psychologists. The significance of our findings suggests this should change.

Part of the issue here, of course, is that this is an emerging field of research. As one of the commentators quoted above noted:

> “we are the research. There is no one before us.”

They ended this quote noting:

> “I think it’s positive news when five months ago we were not taken seriously and now we are”

Of course, smell and taste loss are not new phenomena and there is a growing body research documenting its effects), but the global COVID-19 pandemic has given this issue increased prominence and importance. There is extensive literature on the impact of sight and hearing loss on quality of life, but much less on the loss of taste, smell or chemesthesis (though see 22, 23, 24). The contribution of our senses of smell, taste and chemesthesis to everyday experience, and the emotional importance of tasting and enjoying what we eat, are often not appreciated until they have gone. It is not just health professionals but to a degree all of us who underestimate the importance of these senses to our quality of life. This explains the strong expression from these online posts of being “on the other side of the looking glass”, “feeling alienated”, trying to explain something that neither friends, family or health professionals could fully understand. We hope that this paper will serve to convey the seriousness of this sensory disruption.

A final word as to methodology. Even before Coronavirus limited the possibility of more traditional face-to-face research, social media platforms such as Facebook and Twitter, and online digital platforms such as Facebook messenger have been recognised for their unique value to health research and knowledge translation (14, 25). The value of this online forum was even more pronounced because of the novelty of the condition under examination. Unlike support sites for better-understood conditions, the Covid-19 smell and taste loss Facebook group was constantly evolving and responding to new needs and questions. This is part of a wider trend in which people with Covid-19 and Long Covid have been instrumental in driving the media and research profile of these conditions. This could be seen in action in the way the Facebook site used in this project operated as two-way communication between researchers, experts and people living with altered taste and smell. What could be seen in real time was the sharing and developing of new knowledge about symptoms and treatments/solutions. As such, the AbScent Facebook group represented a novel, rigorous grassroots led model of co-produced, participatory research and knowledge translation in addition to being an important support site for many. This adds further complexity to the growing, but contested, literature on co-produced or user-led research in which an “expert laity” contribute in differing ways (and with more or less radical motivations) to shaping, doing and producing research (15, 26, 27). The active creation of health information and gradual recognition amongst participants that there was no expert bar them on this particular condition led to the appreciation that “we are the research”. This spirit also gives the site an ultimately hopeful, encouraging and salutary ethos, with participants sharing their hard-won stories, tips and hints with each other. This hearing and telling of stories are what brings meaning and order to the chaos of sensory disruption. This form of research could be progressed by equipping participants to contribute to research as citizen scientists (28, 29). This future research could involve participants in codifying these grassroots resources to make them available to clinicians and the wider public.

A limitation of this exploratory research is that it very much constitutes a “snapshot” of the impact of smell and taste loss on well being. For ethical and methodological reasons, we could not trace individual illness trajectories over time and relied solely on the information given to us in response to our prompt questions. As such we also did not systematically collect data on age or gender. We did not confirm if each one of our participants had a COVID-19 positive test, but we believe this to be of minor importance as the participants clearly did have sudden onset smell and taste loss which is now agreed to be strongly indicative of COVID-19 infection.

## Conclusion

The impact of Covid-19 on the senses cannot be viewed as a mild effect, particularly given the impacts may last for months. Covid-19 related sensory upheaval has serious implications for food, eating, health, work and well-being and for some is a profound existential assault disturbing their relationship to self, others and the world. Health care professionals often overlook these serious consequences of smell and taste loss, and intervention focus tends to be on dietary change and the olfactory detection of immediate danger (smoke etc). While we do not dispute the seriousness of these concerns, the ways in which they are currently framed in the literature do not accurately present the extent of potential dietary complications in terms of nutritional compromise, weight loss and weight gain shown above. Our research suggests that the impact of smell and tastes alteration is far reaching and concerning, and that any future research or interventions need to take this broader impact of sensory disruption into consideration.

## Data Availability

The data that support the findings of this study are available on request from the corresponding author, [DBW]. The data are not publicly available due to their containing information that could compromise the privacy of research participants.

